# The Anorexia Nervosa Care Pathway: Closing the Treatment Gap to Improve Care for Patients Admitted to Medical Hospitals

**DOI:** 10.1101/2021.06.14.21258873

**Authors:** Kathryn Allen, Kate O Brien, Marese K O’Reilly, Deirbhile Henderson, Aoibhlinn O Toole, Siobhan MacHale, Karen Boland

## Abstract

**Introduction:** Medical complications of malnutrition and refeeding account for approximately half of deaths in anorexia nervosa (AN). The AN Care Pathway (ANCP) was introduced at our institution in 2016 to improve quality of care of patients admitted for medical observation and management. We report results from our review of medical complications and report the impact and adoption of the ANCP.

**Methods:** The ANCP was developed in response to a need to improve quality of medical monitoring of patients with severe AN using Squire Guidelines and the Plan-Do-Study-Act cycle. All patients admitted to a medical hospital with AN between 2010-2020 were included after hospital inpatient enquiry and medical records were reviewed. Descriptive statistics were calculated using Stata (Statcorp).

**Results:** Fifty-one patients (63 admissions) were included. Median BMI was 13.8 kg/m (11.9-22.5). After ANCP implementation in 2016, compliance with recommended daily ECG, thiamine and blood tests improved from 30% (n=8/27) to 86% (n=21/36). We report a high rate of medical complications of severe AN including anaemia (n=24, 47%), neutropoenia (n=18, 35%), abnormal liver bloods (n=15, 29%) and half developed refeeding syndrome. One-third patients had cardiovascular compromise including reduced cardiac contractility (n=13, 25%), pericardial effusion (n=7, 14%) and one death. Low BMI was associated with cardiovascular complications (mean BMI 13.5 kg/m vs 15.5 kg/m, p=0.01) and neutropoenia (mean BMI 13.4 kg/m vs 15.4 kg/m, p=0.02).

**Conclusion:** Introduction of the ANCP improved quality of care during medical stabilisation. We report a high rate of medical complications of severe AN in patients admitted to a medical hospital. Use of multidisciplinary care protocols may contribute to quality improvement and improved consistency of care for this vulnerable population.

## Introduction

Anorexia nervosa (AN) is an eating disorder which disproportionately affects females and is characterised by weight loss or difficulty maintaining appropriate body weight and distorted body image. DSM-5 criteria for diagnosis include restriction of energy intake relative to requirement leading to low body weight, fear of gaining weight, disturbance in how body weight or shape is experienced or denial of the seriousness of current low body weight. Risk of serious morbidity and mortality associated with AN is significantly greater in this cohort than other female inpatients with psychiatric illness with all-cause mortality of 5.9% or 0.6% per year ^1^. Alarmingly, the mortality rate for young women age 15-24 with AN is over 12 times greater than the unaffected general population ^1^ with the greatest risk during the first 10 years of follow-up after diagnosis ^2^.

Access to specialist eating disorder services in Ireland varies depending on area of residence. In 2018 a blueprint for a National Model of Care for Eating Disorders was published by the Health Service Executive., aiming to address the absence of sufficient eating disorder teams for outpatient care of these vulnerable patients. The current model of care mandates that a proportion of patients with severe malnutrition and/or physical complications of malnutrition require admission to a medical hospital for observation during the acute high-risk phase of feeding, and especially if there are additional medical co-morbidities including diabetes mellitus ^3^. These patients with severe AN and critically low BMI are assessed and monitored in acute medical settings, often without direct access to specialist eating disorder services. In order to standardise and improve the quality of medical monitoring among these at-risk patients, in 2016 a formal protocol was developed and implemented at our institution for identification and care of patients with eating disorders requiring medical admission. This was modelled on MARSIPAN (Management of really sick patients with anorexia nervosa) which outlines recommendations for admission to medical hospital in patients who require medical treatment not available in the acute psychiatric setting as well as the recommended healthcare professionals who should be involved in patient care ^4^. The Anorexia Nervosa Care Pathway (ANCP) has clear parameters assisting emergency, general medical teams and general practitioners referring patients for medical observation and stabilisation (Supplementary Figure 1). Our aim was to address the gap in effective therapy for patients with eating disorders in the acute medical setting ^5^ by providing a model of care that aimed to use what was known about effective treatment to deliver evidence based consistent care. We performed a quantitative assessment of compliance with the ANCP. As a secondary aim, we analysed the rate of medical complications in patients with severe AN admitted for refeeding in this retrospective observational study and potential associations between medical complications of AN and body mass index (BMI) at admission.

## Materials and Methods

### Participants and data collection

A retrospective review of patients admitted primarily for management of AN was performed at Beaumont Hospital, Ireland. Diagnosis of AN was made by consultant psychiatry services using DSM-5 criteria. Patients admitted for other causes with a coincidental history of AN were excluded. Ethical approval for this service evaluation was granted by the local Research Ethics Committee. Hospital Inpatient Enquiry (HIPE) data was requested for inpatients with a discharge diagnosis of AN from January 2010 to May 2020. Medical records were reviewed to confirm diagnosis, indication for admission and investigations and management in keeping with the approved care pathway. Patients admitted for other causes with stable AN were excluded if they did not require refeeding or inclusion on the ANCP. Patient-specific demographic data and hospital admission data were recorded. Medical complications at admission but also those that developed over the course of admission as a complication of the refeeding syndrome (at least 24 hours after admission) were recorded including anaemia (haemoglobin < 11g/dl, neutropoenia < 1.5×10^9^/l, abnormal liver blood tests, cardiovascular complications including pericardial effusion, bradycardia, arrhythmia and impaired ejection fraction, and electrolyte abnormalities including hyponatraemia, hypophosphatemia, hypokalaemia, hypomagnesaemia and hypocalcaemia).

### Anorexia Nervosa Care Pathway

The Anorexia Nervosa Care Pathway (ANCP (supplementary figure 1) was developed based on MARSIPAN (Management of really sick patients with anorexia nervosa) guidelines with formation of a multidisciplinary team for management of patients with severe AN. The ANCP was developed as part of the Plan-Do-Study-Act (PDSA) cycle^6^ with use of the SQUIRE Guideline checklist ^7^. Our unit is a general gastroenterology unit within an acute medical hospital, not a designated site for the provision of specialist eating disorder services. The protocol provides guidance on the management of safe refeeding in this at-risk cohort and early identification of medical complications and hypoglycaemia. In addition, psychological supports and where appropriate, social supports are provided to the patient by the liaison psychiatry and social care team. Multidisciplinary team members include a Gastroenterologist, Liaison Psychiatrist, senior dietitian and clinical nurse manager. Refeeding and treatment regimens were agreed, and progress reviewed at multidisciplinary team meetings with the patient in parallel with planning for transfer to specialist eating disorder services when stable or BMI > 14.5 kg/m. Compliance with key components of the ANCP (supplementary figure 1) was recorded as well as length of stay, readmission rate and medical complications before (2010-2015) and after (2016-2020) implementation.

### Statistical analysis

Descriptive statistics were analysed and reported as median and range. Outcome analysis was performed with linear regression, correcting for age and gender. Data were considered statistically significant if p<0.05. Analysis was performed using Stata (Stata Corp).

## Results

### Description of patient cohort

Medical records of 89 patients coded as a discharge diagnosis of AN between 2010 and 2020 were reviewed. We noted an increasing trend over time of patients admitted to the hospital with a diagnosis of AN as either the primary cause of admission or as a comorbidity. Fifty-one patients with total 63 admissions were identified for inclusion and were admitted for medical observation and stabilisation during feeding. The majority were female (n=49, 96%) with median age of 25 years. Markers of nutritional status included admission weight (median 37.9 kg) and BMI (13.8 kg/m). Eight patients had 2 or more admissions for feeding and medical stabilisation during the study period. Additional nutrition supports were prescribed as nasogastric feeding in 47% (n=24). Most patients were discharged to home with community-based psychiatry follow-up (n=35, 67%). Alternative discharge destinations included specialist eating disorder units in Ireland or the United Kingdom (n=11, 21.5%), other acute medical hospitals (n=2, 4%), psychiatric facilities (n=4, 8%), nursing homes (n=1, 2%) and we unfortunately report one death (Table 1).

**Table 1:** Descriptive table of patient characteristics and metrics of hospital stay subsequent care after discharge.

| Variable | All patients (n=51) | 2010-2015 (n=24) | 2016-2018 (n=27) |
| --- | --- | --- | --- |
| Female | 49 (96%) | 23 (96%) | 26 (96%) |
| Age (median, range years) | 25 (16-59) | 25 (17-59) | 24 (16-51) |
| Length of Stay (days) | 12 | 8.5 (2-87) | 15.5 (2-188) |
| Readmissions | 9 (18%) | 5 | 4 |
| BMI at admission (kg/m<sup>2</sup>) | 13.8 | 14.4 | 13.3 |
| Average weight gain (% of admission weight) | 8.6% | 8.63% | 8.66% |
| Nasogastric feeding | 24 (47%) | 11 (49%) | 13 (48%) |
| Discharge destination: Home | 35 (67%) | 18 (75%) | 17 (63%) |
| Convalescence | 1 (2%) | 1 (4%) | 0 |
| Specialist Unit | 11 (21.5%) | 5 (21%) | 6 (22%) |
| Medical Hospital | 2 (4%) | 0 | 2 (8%) |
| Psychiatric Hospital | 4 (8%) | 0 | 4 (15%) |

### Medical complications during acute nutrition support intervention in anorexia nervosa

Incidence of medical complications were recorded through review of medical, radiology and laboratory records. Almost half (47%, n=24) had anaemia on presentation, attributed to acute upper gastrointestinal haemorrhage and peptic ulcer disease in 3 patients (6%, Figure 1A). Eighteen (35%) patients had neutropoenia (<1.5×10^9^/l), severe (<1 x10^9^/l) in half of these patients. None had clinically significant infection or septicaemia, although three were treated with granulocyte-colony stimulating factor (G-CSF) due to critically low neutrophil counts. Presence of neutropoenia was associated with lower mean BMI (13.4 kg/m vs 15.4 kg/m, p=0.02, CI -3.6669 to -0.3605).

**Figure 1:**
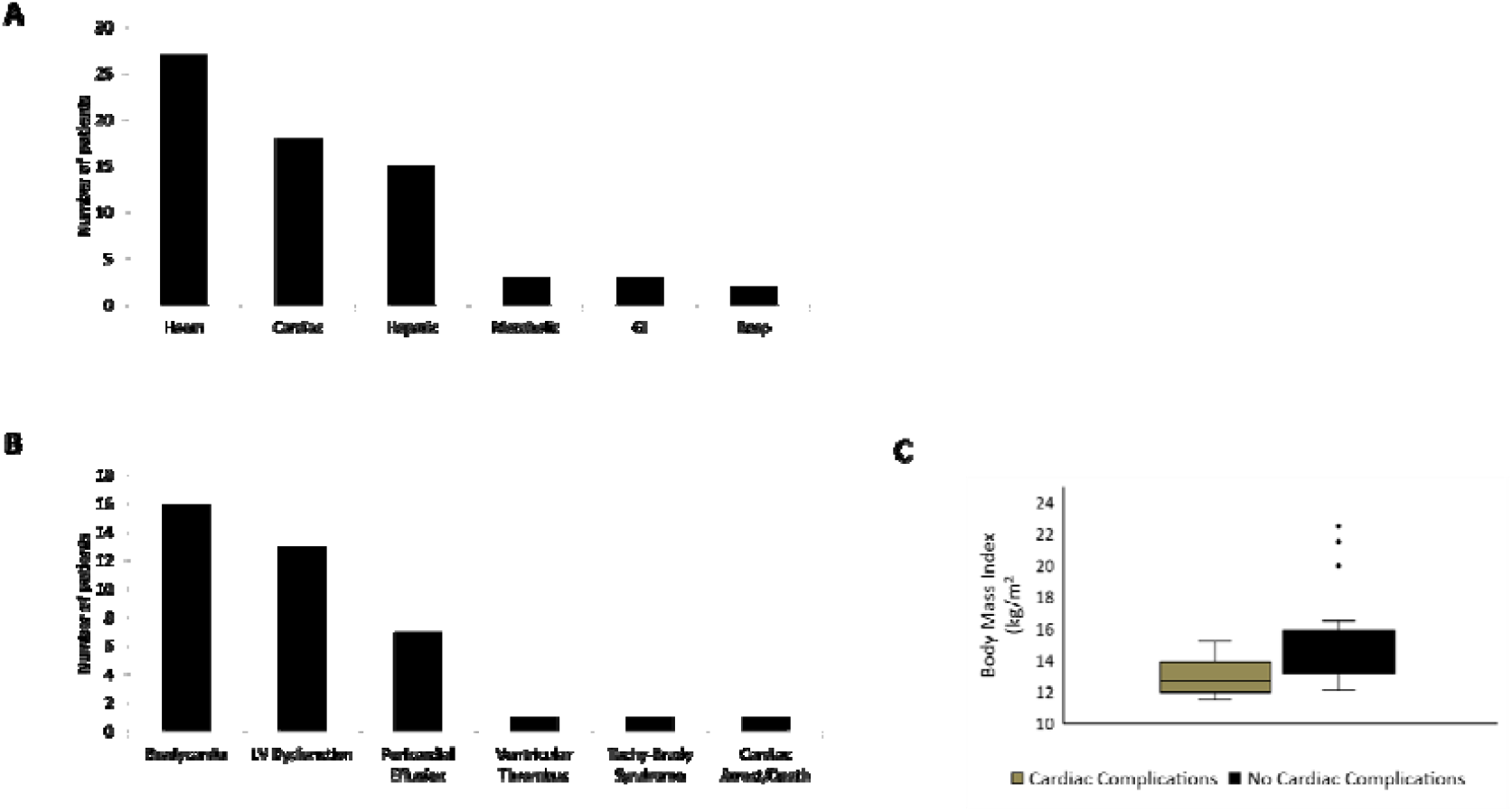
Medical Complications of inpatients with anorexia nervosa. **1A:** Number of patients admitted with medical complications recorded during their inpatient stay. **1B**: Cardiovascular complications of inpatients with anorexia nervosa. **1C:** Data depict mean BMI in patients with cardiovascular complications, * p=0.01

Other abnormal blood investigations included abnormal liver blood tests (n=15, 29%) and refeeding syndrome (n=25, 49%). Manifest refeeding syndrome (biochemical shifts with pedal oedema, tachypnoea, cardiac or neurological disturbance) was diagnosed in 25% (n=13) patients ^8 9^. Risk of the refeeding syndrome did not correlate with low weight or BMI (p=0.8). Additional complications included two patients who presented with diabetic ketoacidosis due to manipulation of diet and insulin regimes with known AN and complications of metabolic bone disease in these inpatients including osteomalacia (n=1) and atraumatic pubic ramus fracture (n=1, Figure 1A).

Cardiovascular complications were common reflecting disease severity among those admitted. Sixteen (31%) patients had persistent bradycardia (heart rate < 50 beats per minute over a period greater than 24 hours), with junctional escape rhythms in 4 without QT prolongation. Patients with profound bradycardia, significant pedal oedema or evidence of cardiac decompensation were referred for transthoracic echocardiogram (TTE). Low ejection fraction and/or ventricular hypokinesis was reported in 13 (25%) patients, and pericardial effusion in 7 patients (14%). Other cardiac manifestations included ventricular thrombus (n=1) and one death in a patient who presented with cardiac arrest due to arrhythmia associated with critical hypokalaemia (Figure 1B). Low BMI on admission was associated with bradycardia or TTE abnormalities with mean admission BMI of 13.5 kg/m compared 15.5 kg/m in patients with cardiovascular symptoms or signs (p=0.01, CI mean -3.4 to -0.45, Figure 1C).

### Introduction of AN Care Pathway and transfer to gastroenterology core-ward care was associated with improved monitoring of medical complications and quality of care

Thirty-six admissions (57%) were recorded after introduction of the ANCP. Cornerstones of the ANCP included transfer of care to the gastroenterology service on speciality-specific ward, daily blood monitoring, thiamine prescription and daily ECG per protocol. In addition, regular multidisciplinary team meetings with the patients at least once weekly were attended by the patient and their family, consultant gastroenterologist, liaison psychiatrist, senior dietitian and the ward’s clinical nurse manager. Following introduction of ANCP, a greater proportion of patients with severe AN (82% vs 46%) were transferred to gastroenterology services for multidisciplinary management.

Before ANCP, 70% admissions did not meet accepted national standard of care for monitoring of the refeeding syndrome and its complications with daily ECG, electrolyte monitoring and thiamine prescription ^8^. This significantly improved after introduction of the ANCP and education around this protocol for nursing and medical professionals. Our evaluation found that only on 5 admissions were these criteria not met after ANCP introduction in 2016 (14%, n=5/36). There is a temporal trend of improved monitoring for refeeding of these patients which accelerated after introduction of the ANCP and associated education (Figure 2).

**Figure 2:**
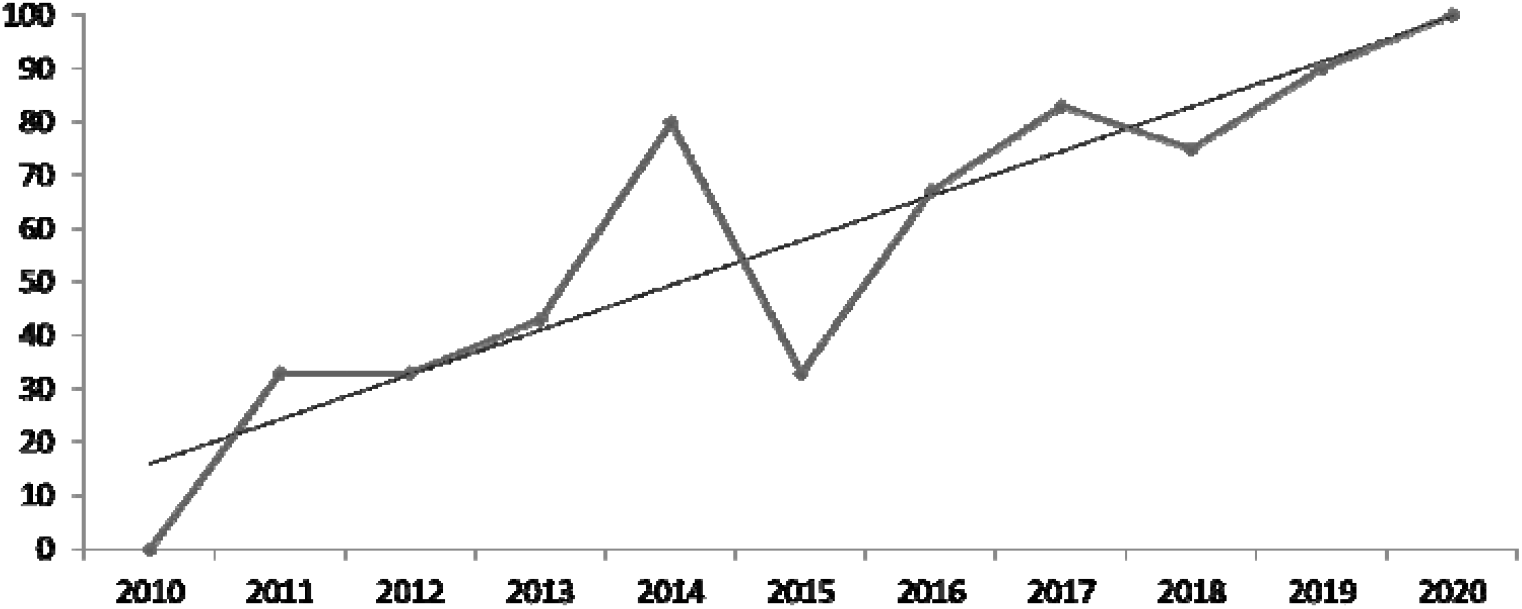
Compliance with Anorexia Nervosa Care Pathway. Data depict proportion of admissions annually compliant with cornerstones of the anorexia nervosa care pathway (referral to dietitian and psychiatry, daily lab monitoring for 5 days, daily ECG for 3 days, thiamine) before and after implementation, and associated trendline for compliance rate.

Full compliance with the complete ANCP was also audited including electrolyte monitoring, prescription of thiamine, electrocardiograph (ECG) monitoring, transfer to gastroenterology specialty ward, referral to liaison psychiatry and dietitians and weekly multidisciplinary patient-centred meetings. Half of admissions for medical stabilisation met these strict criteria for full compliance with best practice measures. Areas identified for ongoing quality improvement included early ECG monitoring and weekly scheduled multidisciplinary team meetings with patients to collaboratively progress and amend treatment plans (68% compliance). However, almost three-quarters of admissions where management did not comply with all aspects of the ANCP occurred in the first 18 months after introduction, showing improvement over time consistent with ongoing education of medical and nursing staff and increased awareness of presence of this protocol on the hospital intranet.

Readmission rate was similar before and after introduction of ANCP. While there was no statistically significant difference in length of stay, mean admission days were greater in the group post ANCP introduction (15.5 vs 8.5 days, p=0.2), which may be related to the fact that patients presenting since 2016 had a lower mean BMI (13.3 kg/m vs 14.4 kg/m) and more medical complications of AN including cardiac decompensation (p=0.04), hence requiring additional time to achieve stabilisation prior to discharge. Patients with severe AN admitted after ANCP were not more likely to be referred to inpatient specialist eating disorder units elsewhere, in keeping with the goal of the Health Service Executive of Ireland to facilitate discharge to home with outpatient specialist multidisciplinary care for these complex patients.

## Discussion

We note a high rate of medical complications among patients with severe AN admitted for medical observation and refeeding. Service evaluation of the anorexia nervosa care pathway (ANCP) determined that cornerstones of monitoring for manifest refeeding syndrome and complications of severe AN were not consistently applied in this cohort before 2016. Implementation of the ANCP care pathway improved and standardised quality of care during the high-risk refeeding period and established an ethos of consistent multidisciplinary communication and management for these vulnerable patients in keeping with the national blueprint for the management of patients with eating disorders.

In 2016, a survey distributed among hospital and community-based medical practitioners in Ireland identified that respondents had poor recognition of the symptoms of eating disorders. Non-psychiatrists were not as confident in their ability to treat patients with eating disorders and associated complications (p<0.0001)), demonstrating the need to improve and build on education relating to these conditions^10^. Kazdin and colleagues have previously discussed the critical need to address the “research-practice gap” in management of mental health and eating disorders, but crucially, addressing the “Treatment Gap”. This refers to the extension of evidence-based treatments for mental health disorders, disseminating evidence-based practice in ways that will reach a large number of patients in a variety of settings ^5 11^. System factors relating to availability of specialist eating disorder services and attitudinal factors relating to physician training and knowledge may impact access to evidence based, quality care. Our goal was to achieve a consistent standard of care for the patients in the inpatient setting which addressed this treatment gap. Our aim was to develop a process for provision of psychological support for patients while receiving medical attention and refeeding, and further to establish a platform to access outpatient specialist eating disorder services. In order to achieve this, we developed the ANCP which is accessible electronically, and facilitated training for medical and nursing practitioners through local ward-based education, hospital-wide Grand Rounds presentations and dissemination of the ANCP through the hospital-run smartphone application. We have demonstrated improvements in quality of care for this vulnerable inpatient group through implementation of the ANCP (Figure 2) and adoption of a policy which includes regular multidisciplinary meetings. The goal of the ANCP is provide best quality care for these patients and a clinical model for multidisciplinary care with input from dietitians, gastroenterologists and liaison psychiatry services with structured discharge to outpatient or inpatient specialist services as appropriate.

Medical admission targets focus on stabilisation and medical management of complications of severe AN with concurrent development of a treatment plan in a setting commensurate with their needs. The ANCP achieved greater consistency and improved quality of care for these patients (Figure 2). Improving compliance over time since 2016 reflects the importance of ongoing training and education of medical and nursing staff to raise aware of the presence and importance of care protocols and pathways. Implementation of the ANCP had no statistically significant impact on length of stay, discharge destination or of readmissions for medical treatment. Evidence consistently points to AN as a prolonged disease with high relapse rate of approximately 31% ^12^ and our protocol was designed to provide a platform for safe care during their medical stay rather than impact long term prognosis relating to their underlying eating disorder.

AN is associated with a heavy burden of medical complications ^13 14^. The magnitude of the risk associated with refeeding for critically malnourished patients with AN admitted for medical observation during this high-risk period may be challenging to interpret and differentiate from patients admitted with symptomatic medical conditions. Sullivan completed a valuable meta-analysis of 42 studies inclusive of >3000 patients which revealed a crude mortality rate of 5.9%, and over half of these death related to medical complications of eating disorders ^1^. In this study, we report one death due to cardiac arrest in a patient admitted with AN. Cardiovascular stress is well recognised in severe AN during feeding and the early phase of nutrition support with 56% patients admitted with extreme AN having bradycardia on admission in one study ^15^. This is, in many cases, related to physiological compensatory mechanisms reducing metabolic demand in severe starvation. In addition, patients with severe AN may exhibit increased parasympathetic tone, reduction in intracardiac glycogen stores and myocardial atrophy with structural changes evidence on cardiac magnetic resonance imaging (MRI) studies ^16 17^. Sixteen (33%) patients in our study had bradycardia with documented evidence of junctional rhythms in a quarter, although none required pacemaker therapy. Pedal oedema, chest pain, syncope or tachy-brady syndrome were indications for transthoracic echocardiogram (TTE). Twenty-nine percent of patients met these criteria, with abnormal findings in 13/16 (81%) of those who had TTE including low ejection fraction and/or hypokinesia, pericardial effusion (n=7/16, 44%) or ventricular thrombus (n=1/16, 6%). At TTE, serum albumin level did not correlate with likelihood of pericardial effusion (p=0.9). However, mean BMI in affected was significantly lower (p=0.01, Figure 1C) consistent with physiological changes associated with cardiovascular stress. There are few studies with data on the prevalence of pericardial effusions in severe AN and these may be more frequent in these patients than is widely recognised ^18^.

Haematological abnormalities have been well described in severe AN as sequelae of malnutrition, often despite normal micronutrient levels. Pancytopoenia in AN is associated with partial or focal gelatinous degeneration of the bone marrow ^19^ which usually normalises with improvement of nutritional status. In one assessment of patients with amenorrhoeic AN, 16.7% patients were anaemic and 7.9% had neutropoenia (<1.5×10^9^/l) ^20^. We noted higher prevalence of anaemia < 10 g/dL (45%, n=24) after exclusion of patients with upper gastrointestinal haemorrhage, in addition to higher rate of neutropoenia (35%, n=18) which was associated with lower mean BMI (p=0.02). There were no recorded cases of septicaemia despite neutropoenia or leucopoenia. The proportion of patients with anaemia was comparable to an analysis of patients with extreme AN admitted for medical stabilisation to a specialist unit ^15^. The higher proportion of anaemia in these cohorts reflects severe disease and lower BMI at the time of treatment.

We describe a single-centre experience of prevalence and management of medical complications of patients with severe AN admitted for medical observation and refeeding. Strengths include the fact that inclusion was limited to patients with severe AN admitted primarily for management of their eating disorder rather than patients admitted for management of other medical conditions with a coincidental diagnosis of AN. The study is limited by patient numbers and retrospective study design. Development of nursing and medical expertise with a strong multidisciplinary focus optimises both medical care of complications and treatment of AN during this acute phase. We report successful implementation of a centre-specific care pathway targeting improved and consistent quality of care for these vulnerable patients, aiming to in part, address the “treatment gap” that exists for management of eating disorders in the acute medical setting, outside specialized eating disorder programmes. Implementation of the ANCP standardised quality of care during the high-risk refeeding period and established an ethos of consistent multidisciplinary management for these vulnerable patients in keeping with the national blueprint for the management of patients with eating disorders.

## Data Availability

Available on request

## References

1. Sullivan PF. Mortality in anorexia nervosa. Am J Psychiatry 1995;152(7):1073–4. doi: 10.1176/ajp.152.7.1073 [published Online First: 1995/07/01]

2. Franko DL, Keshaviah A, Eddy KT, et al. A longitudinal investigation of mortality in anorexia nervosa and bulimia nervosa. Am J Psychiatry 2013;170(8):917–25. doi: 10.1176/appi.ajp.2013.12070868

3. HSE BotwotnwgatcagotCoPiI. Eating Disorder Services. HSE Model of Care for Ireland. 1st ed. Dublin, 2017.

4. MARSIPAN TRCoP, Physicians and Pathologists. MARSIPAN: Management of Really Sick Patients with Anorexia Nervosa. CR189. 2nd ed. London, 2014.

5. Kazdin AE, Fitzsimmons-Craft EE, Wilfley DE. Addressing critical gaps in the treatment of eating disorders. Int J Eat Disord 2017;50(3):170–89. doi: 10.1002/eat.22670 [published Online First: 2017/01/20]

6. Taylor MJ, McNicholas C, Nicolay C, et al. Systematic review of the application of the plan– do–study–act method to improve quality in healthcare. BMJ Quality & Safety 2014;23(4):290–98. doi: 10.1136/bmjqs-2013-001862

7. Ogrinc G, Davies L, Goodman D, et al. SQUIRE 2.0 (Standards for QUality Improvement Reporting Excellence): revised publication guidelines from a detailed consensus process. BMJ Qual Saf 2016;25(12):986–92. doi: 10.1136/bmjqs-2015-004411 [published Online First: 2015/09/16]

8. Karen Boland Coh, Damodar Solanki. Prevention and Treatment of Refeeding Syndrome in the Acute Care Setting. 1 ed. Dublin: Irish Society for Clinical Nutrition and Metabolism, 2014.

9. Singer P, Blaser AR, Berger MM, et al. ESPEN guideline on clinical nutrition in the intensive care unit. Clin Nutr 2019;38(1):48–79. doi: 10.1016/j.clnu.2018.08.037 [published Online First: 2018/10/24]

10. McNicholas F, O’Connor C, O’Hara L, et al. Stigma and treatment of eating disorders in Ireland: healthcare professionals’ knowledge and attitudes. Irish Journal of Psychological Medicine 2016;33(1):21–31. doi: 10.1017/ipm.2015.24 [published Online First: 2015/05/22]

11. Kazdin AE. Addressing the treatment gap: A key challenge for extending evidence-based psychosocial interventions. Behaviour Research and Therapy 2017;88:7-18. doi: https://doi.org/10.1016/j.brat.2016.06.004

12. Berends T, Boonstra N, van Elburg A. Relapse in anorexia nervosa: a systematic review and meta-analysis. Curr Opin Psychiatry 2018;31(6):445–55. doi: 10.1097/yco.0000000000000453 [published Online First: 2018/08/17]

13. Gibson D, Workman C, Mehler PS. Medical Complications of Anorexia Nervosa and Bulimia Nervosa. Psychiatr Clin North Am 2019;42(2):263–74. doi: 10.1016/j.psc.2019.01.009 [published Online First: 2019/05/03]

14. Westmoreland P, Krantz MJ, Mehler PS. Medical Complications of Anorexia Nervosa and Bulimia. Am J Med 2016;129(1):30–7. doi: 10.1016/j.amjmed.2015.06.031 [published Online First: 2015/07/15]

15. Gibson D, Watters A, Cost J, et al. Extreme anorexia nervosa: medical findings, outcomes, and inferences from a retrospective cohort. Journal of Eating Disorders 2020;8(1):25. doi: 10.1186/s40337-020-00303-6

16. Oflaz S, Yucel B, Oz F, et al. Assessment of myocardial damage by cardiac MRI in patients with anorexia nervosa. Int J Eat Disord 2013;46(8):862–6. doi: 10.1002/eat.22170 [published Online First: 2013/08/08]

17. Yahalom M, Spitz M, Sandler L, et al. The significance of bradycardia in anorexia nervosa. Int J Angiol 2013;22(2):83–94. doi: 10.1055/s-0033-1334138 [published Online First: 2014/01/18]

18. Frölich J, von Gontard A, Lehmkuhl G, et al. Pericardial effusions in anorexia nervosa. Eur Child Adolesc Psychiatry 2001;10(1):54–7. doi: 10.1007/s007870170047 [published Online First: 2001/04/24]

19. Abella E, Feliu E, Granada I, et al. Bone marrow changes in anorexia nervosa are correlated with the amount of weight loss and not with other clinical findings. Am J Clin Pathol 2002;118(4):582–8. doi: 10.1309/2y7x-ydxk-006b-xlt2 [published Online First: 2002/10/12]

20. De Filippo E, Marra M, Alfinito F, et al. Hematological complications in anorexia nervosa. Eur J Clin Nutr 2016;70(11):1305–08. doi: 10.1038/ejcn.2016.115 [published Online First: 2016/11/03]

